# Clinical judgement of General Practitioners for the diagnosis of dementia: a diagnostic test accuracy study

**DOI:** 10.1101/2020.11.20.20234062

**Authors:** Samuel Thomas Creavin, Judy Haworth, Mark Fish, Sarah Cullum, Antony Bayer, Sarah Purdy, Yoav Ben-Shlomo

## Abstract

**Background:** The accuracy of General Practitioners’ (GPs’) clinical judgement for dementia is uncertain.

**Aim:** Investigate the accuracy of GPs’ clinical judgement for the diagnosis of dementia.

**Design and Setting:** Diagnostic test accuracy study, recruiting from 21 practices around Bristol.

**Method:** The clinical judgement of the treating GP (index test) was based on the information immediately available at their initial consultation with a person aged over 70 years who had cognitive symptoms. The reference standard was an assessment by a specialist clinician, based on a standardised clinical examination and made according to ICD-10 criteria for dementia.

**Results:** 240 people were recruited, with a median age of 80 years (IQR 75 to 84 years), of whom 126 (53%) were men and 132 (55%) had dementia. The median duration of symptoms was 24 months (IQR 12 to 36 months) and the median ACE-III score was 75 (IQR 65 to 87). GP clinical judgement had sensitivity 56% (95% CI 47% to 65%) and specificity 89% (95% CI 81% to 94%). Positive likelihood ratio was higher in people aged 70-79 years (6.5, 95% CI 2.9 to 15) compared to people aged ≥ 80 years (3.6, 95% CI 1.7 to 7.6), and in women (10.4, 95% CI 3.4 to 31.7) compared to men (3.2, 95% CI 1.7 to 6.2), whereas the negative likelihood ratio was similar in all groups.

**Conclusion:** A GP clinical judgement of dementia is specific, but confirmatory testing is needed for symptomatic people who GPs judge as not having dementia.

**How this fits in:** Previous studies in this area have investigated the accuracy of GP clinical judgement as a screening test for dementia in unselected people attending a primary care clinic; or as a retrospective test based on their knowledge of their patient; or derived the accuracy of judgement from the medical records, which may not reflect the judgement of the clinician. The role of the GP in supporting a more effective route to diagnosis for people with dementia is a research priority for patients, carers and clinicians. This study shows that, in a symptomatic older adult, prospective clinical judgement may be useful for helping to confirm a diagnosis of dementia, whereas GP judgement should not by itself be used to exclude dementia.

## Introduction

The James Lind Alliance has identified the role of general practice in supporting a more effective route to diagnosis of dementia as a priority for health research (1). People with symptoms of dementia have historically faced long delays to get an assessment and an explanation for their symptoms (2). Approaches to address waiting lists have included psychiatrists supporting primary care memory clinics (3), integrated one-stop clinics (4), and training GPs to make a diagnosis in uncomplicated cases (5, 6) which is supported by NICE (7). Some GPs have in the past been hesitant about diagnosing dementia when there is no disease modifying treatment (8). A GP could use a range of brief cognitive assessments (9) to evaluate a person with symptoms of dementia. National guidelines differ on which test to use, possibly because there is little evidence in a symptomatic primary care population (10, 11). Formally evaluating cognition takes time, and familiarity with the test. GPs report using non-standardised processes (12), such as their clinical judgement (13) to decide whether a person has dementia. The sensitivity of GP clinical judgement for diagnosing dementia has been reported between from 51% (14) to 100% (15), and the specificity ranges from 58% (16) to 100% (15). Previous studies to investigate the accuracy of GP clinical judgement have typically suffered from one of two significant limitations (17). Firstly, a definition of clinical judgement which is of unclear relevance to practice, such as retrospective judgement, or indeed documentation of recorded diagnoses in the medical record, which are systematically incomplete (18). Secondly, sampling unselected people attending general practice regardless of symptoms, which is more akin to screening. To address these limitations, we investigated the prospective accuracy of GP clinical judgement for the diagnosis of dementia syndrome in people over 70 years (19).

## Methods

### Population

We recruited participants from 21 participating GP surgeries in the Bristol, North Somerset, and South Gloucestershire (BNSSG) area, which is a diverse geographic area within 15 miles of the City of Bristol, covering a total population of around 900,000 people across 82 GP practices. Research clinics were in four participating GP surgeries, strategically located for accessibility. We calculated that a minimum sample size of 200 was needed for a lower bound of the specificity 95% confidence interval of 80%, based on a specificity of 95% in prior studies, and a 75% prevalence of dementia in local memory clinic data (20).

### Inclusion and exclusion criteria

Participants were people with cognitive symptoms but no prior diagnosis of dementia, who were aged at least 70 years and had been referred by their GP to this research study. Cognitive symptoms were not specified but generally include disturbance in memory, language, executive function, behaviour, and visuospatial skills (21). Symptoms were required to be present for at least six months, and could be reported by the person themselves, a family member, a professional, or another person; there was no severity threshold. Cognitive problems did not need to be the main focus of the consultation. As is routine practice, GPs could initiate an enquiry about cognition if they perceived there was a problem. Symptom duration was determined from the clinical history. An accompanying informant was mandatory. All participants were offered free accessible transport and translation services. People were excluded if they had a known neurological disorder (i.e. Parkinsonism, Multiple Sclerosis, learning disability, Huntington’s disease), registered blind, profound deafness (i.e. unable to use a telephone), psychiatric disorder requiring current secondary care input, or if cognitive symptoms were either rapidly progressive or co-incident with neurological disturbance. People with cognitive problems that were so advanced that they were unable to consent were excluded as they were judged by a lay advisory group to find the research process overly burdensome. GPs were encouraged to make a clinical judgement and refer a consecutive series of all eligible patients with cognitive symptoms to the study, regardless of what their clinical judgement was or any test results. GPs gave study information including a leaflet, and obtained verbal consent to share contact details with the study on a referral form, including the persons age, sex, contact details, and the GPs clinical judgement. The study team contacted people referred by GPs to re-confirm eligibility, provide further written study details, and offer a research clinic appointment. The research team took written consent from all participants.

### Index test of clinical judgement

The referring GP recorded their clinical judgement using an electronic referral form during a consultation with their patient about cognitive symptoms. Clinical judgement was operationalised as normal, cognitive impairment not dementia (CIND), or dementia as options for response to the question *“Is your gut feeling that this person”*. GPs were not required to arrange any test and could also refer people simultaneously or subsequently to NHS services. The study team contacted the practice at least three times to obtain any missing referral data.

### Reference standard

At the research clinic, a single specialist physician with more than 20 years’ experience in the field of dementia conducted a standardised assessment lasting approximately 60 minutes comprising clinical history, the Addenbrooke’s Cognitive Examination III (ACE-III (22), Brief Assessment Schedule Depression Cards (BAS-DEC) (23) and the Bristol Activities of Daily Living (BADL) Questionnaire (24). The specialist was not aware of other test results such as GP judgement or any investigations. The reference standard was based on the evaluation of the specialist physician for dementia according to ICD-10 criteria (25). Medical records were reviewed for all participants six months after the research clinic to identify any information that had come to light that would contradict this judgement. A second specialist adjudicated cases where there was diagnostic uncertainty at the research clinic using the initial specialist assessment and the medical record review, but without access to the GP judgement. Study data were electronically entered and managed using RED-Cap (Research Electronic Data Capture) hosted at the University of Bristol (26).

### Statistical methods

Characteristics of participants including age, sex and ACE-III score were tabulated by dementia status according to the reference standard. Separate logistic regression analyses were used with non-participation as the dependent variable and GP judgement, age (in years) and female sex as the independent variables to test the hypothesis of no association with these variables. Time from referral to appointment was described using median and interquartile range and logistic regression was used to test the hypothesis of no association between time to appointment (in days) and dementia (as the dependent variable). Measures of diagnostic test accuracy (sensitivity, specificity, positive and negative likelihood ratios (LRP, LRN), predictive values) were calculated together with 95% confidence intervals, for GP judgement of dementia against reference standard of dementia. Decision curve analysis (27) was used to show the net benefit (incorporating discrimination and calibration) of GP judgement at varying threshold probabilities (P_*t*_). Decision curve analysis quantifies the net benefit of a test, in units of true positive, across a range of preferences (28) where a net benefit of 0.05 means “five true positives for every 100 patients in the target population” (29), compared to the “label:all” (worried about missing dementia) or “label:none” (worried about overdiagnosis). The net benefit equation incorporates true (TP) and false positives (FP), but not test negatives (30):

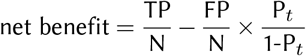

Sensitivity analyses were done to explore whether accuracy varied by age (<80 years | ≥ 80 years) since prediction models perform differently in these age groups (31), and sex. Cochran’s Q test was used to test the hypothesis of no difference in likelihood ratios between groups (32).

## Results

### Participants

Recruitment took place between March 2015 and May 2017. Figure 1 shows a flowchart for inclusion in the study. The theoretically “eligible” figure of 1,735 people was derived from the age specific incidence of dementia (33) and the demographics of the population in the participating practices (34,956 aged over 70 years (34). The number approached is unknown. One person who consented withdrew before any data collection was done because they were admitted acutely ill to hospital. Of the 240 with available data, there were 20 borderline cases that were adjudicated by a second specialist. The 240 people were classified by the reference standard as either Normal (47), Dementia (132) of whom 1 had DSM-5 but not ICD-10 because they had subjective but not objective amnesia, or were CIND (61) of 59 whom met criteria for MCI (1 affective disorder, 1 brain injury). Compared to people who participated, there was little evidence of an association between non-participation and a GP clinical judgement of CIND (odds ratio 1.2; 95% CI 0.55 to 2.41) or dementia (odds ratio 1.9; 95% CI 0.90 to 3.93). Compared to people who participated, non-participants were older (odds ratio per year 1.08; 95% CI 1.04 to 1.12), or more often female (odds ratio 1.88; 95% CI 1.21 to 2.92). The median time between referral (clinical judgement) and the clinic appointment (reference standard) was 47 days (IQR 30 to 72 days), the longest interval was 177 days, due to difficulties attending earlier appointments. There was no association between time from referral to appointment and dementia (odds ratio per day 1.0; 95% CI 0.99 to 1.01). Table 1 shows the demographics of participants.

**Table 1.** Characteristics of participants by cognitive category

| Characteristic | Cognitive category * |  | Normal<br>n=47 |
| --- | --- | --- | --- |
|  | Dementia<br>n=132 | CIND<br>n=61 |  |
| **Sex n (column %)** |  |  |  |
| Male | 68 (52) | 35 (57) | 23 (49) |
| Female | 64 (49) | 26 (43) | 24 (51) |
| **Age (years) Median (IQR)** |  |  |  |
| At clinic | 82 (77-86) | 80 (75-83) | 75 (72-80) |
| Left school | 15 (15-16) | 15 (15-16) | 16 (15-16) |
| Retired | 60 (58-65) | 60 (58-67) | 61 (58-65) |
| **Symptom onset** |  |  |  |
| Median (IQR) (months) |  |  |  |
| Time ago | 24 (12-36) | 18 (12-24) | 21 (12-36) |
| Type, n (column %) |  |  |  |
| Gradual | 111 (84) | 55 (90) | 43 (91) |
| Sudden | 13 (10) | 5 (8) | 0 (-) |
| Uncertain | 8 (6) | 1 (1) | 4 (9) |
| **Symptom pattern n (column %)** |  |  |  |
| Course |  |  |  |
| Progressive | 111 (84) | 42 (69) | 29 (62) |
| Stepwise | 2 (2) | 0 (-) | 0 (-) |
| Regressive | 1 (1) | 1 (2) | 1 (2) |
| Static | 5 (4) | 7 (11) | 9 (19) |
| Uncertain | 13 (10) | 11 (18) | 8 (17) |
| **Fluctuation** |  |  |  |
| None | 112 (85) | 53 (87) | 45 (96) |
| Within one day | 12 (9) | 5 (8) | 1 (2) |
| Over several days | 8 (6) | 3 (5) | 1 (2) |
| **ACE-III Score median (IQR)** |  |  |  |
| Total (max 100) | 69 (61-74) | 82 (76-87) | 93 (90-95) |
| **GP opinion n (row %)** |  |  |  |
| Normal | 6 (18) | 9 (26) | 19 (56) |
| CIND | 52 (43) | 41 (34) | 27 (23) |
| Dementia | 74 (86) | 11 (13) | 1 (1) |

Dementia according to ICD-10 definition; MCI according to Petersen definition.
ACE-III Addenbrookes' Cognitive Examination III; CIND Cognitive impairment, not dementia.

**Fig. 1.**
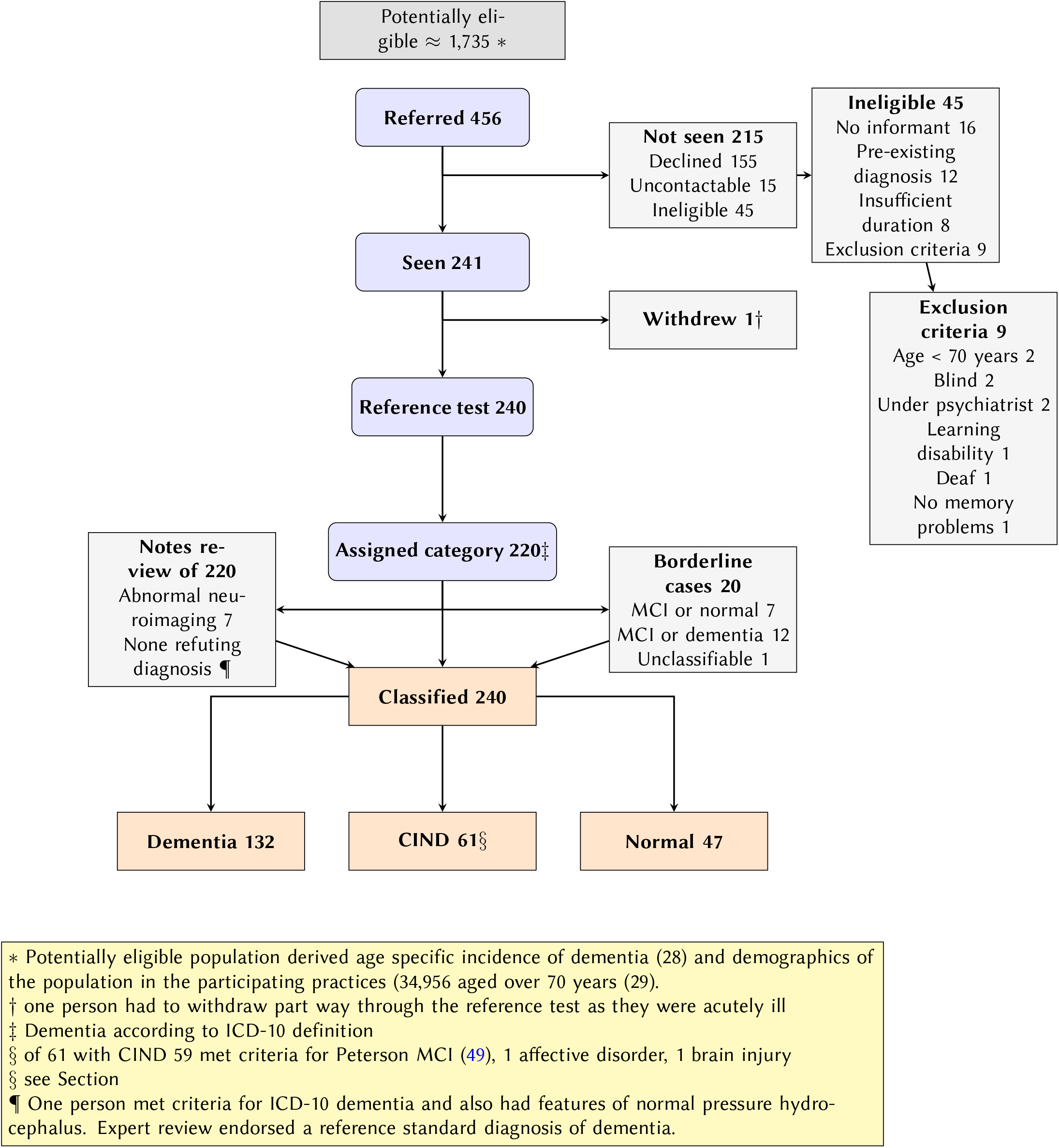
STARDdem flowchart for inclusion of participants in the study

Two people could not complete the ACE-III because English was not their first language; they had both declined an interpreter. In both cases sufficient information was available from other parts of the assessment for a categorisation about cognition to be made (one had normal cognition, one had dementia). For the 238 people who had an ACE-III score, the median was 75 (interquartile range 65 to 87). Referring GPs judged that 34 people were normal, 86 had dementia, and 120 were CIND; the one person who withdrew from the study due to acute illness was judged by the referring GP to have CIND. People that GPs judged as having dementia had a total ACE-III score IQR of 60 to 74, with a 90th centile of 81/100 and a highest score of 95/100. Similarly, people that GPs judged as having CIND had an ACE-III score IQR 71 to 89.

### Diagnostic accuracy

Table 2 shows the diagnostic accuracy for GP judgement for dementia. The sensitivity and specificity of GP judgement were respectively 56% (95% CI 47% to 65%) and 89% (95% CI 81% to 94%). Clinical judgement was more useful for ruling in dementia, than ruling it out, with higher specificity and positive predictive value than sensitivity and negative predictive value. In people aged 80 or more years, clinical judgement had similar sensitivity and specificity to those aged under 80 years (LRP p=0.296 LRN p=0.798). There was weak evidence that clinical judgement in women had a higher LRP (p=0.074) and a lower LRN (p=0.064) than clinical judgement in men.

**Table 2.** Accuracy of GP judgement for the diagnosis of dementia

|  | **Sensitivity**<br>(95% CI) | **Specificity**<br>(95% CI) | **PPV**<br>(95% CI) | **NPV**<br>(95% CI) | **LRP**<br>(95% CI) | **LRN**<br>(95% CI) |
| --- | --- | --- | --- | --- | --- | --- |
| **GP judgement**<br>(n=240) | 56 (47 to 65) | 89 (81 to 94) | 86 (77 to 93) | 62 (54 to 70) | 5.1 (2.9 to 8.8) | 0.49 (0.40 to 0.61) |
| **Age ≥ 80 years**<br>(n=123) | 57 (45 to 67) | 84 (67 to 94) | 89 (77 to 96) | 46 (34 to 59) | 3.6 (1.7 to 7.6) | 0.52 (0.39 to 0.68) |
| **Age < 80 years**<br>(n=117) | 55 (40 to 70) | 91 (82 to 97) | 81 (64 to 93) | 75 (65 to 84) | 6.5 (2.9 to 15) | 0.49 (0.35 to 0.68) |
| **Men**<br>(n= 126) | 50 (38 to 62) | 85 (73 to 93) | 79 (64 to 90) | 59 (48 to 70) | 3.2 (1.7 to 6.2) | 0.59 (0.46 to 0.77) |
| **Women**<br>(n= 114) | 63 (50 to 74) | 94 (84 to 99) | 93 (81 to 99) | 66 (54 to 77) | 10.4 (3.4 to 31.7) | 0.40 (0.29 to 0.55) |

LRN negative likelihood ratio LRP positive likelihood ratio NPV negative predictive value PPV positive predictive value

Figure 2 shows that clinical judgement has greater net benefit than a label:all approach at threshold probabilities of above 50%, and a label:none approach at threshold probabilities below 85%. At a threshold probability of 80%, indicating a preference for avoiding over-diagnosis, clinical judgement has a net benefit of 0.11 over the label:none approach, indicating an additional 11 true positives for every 100 people. If the doctor prefers to not miss dementia, with a threshold probability of 33%, then clinical judgement identifies five fewer true positives for every 100 people than the label:all approach (everyone is treated as if they have dementia: further tests or referral arranged).

**Fig. 2.**
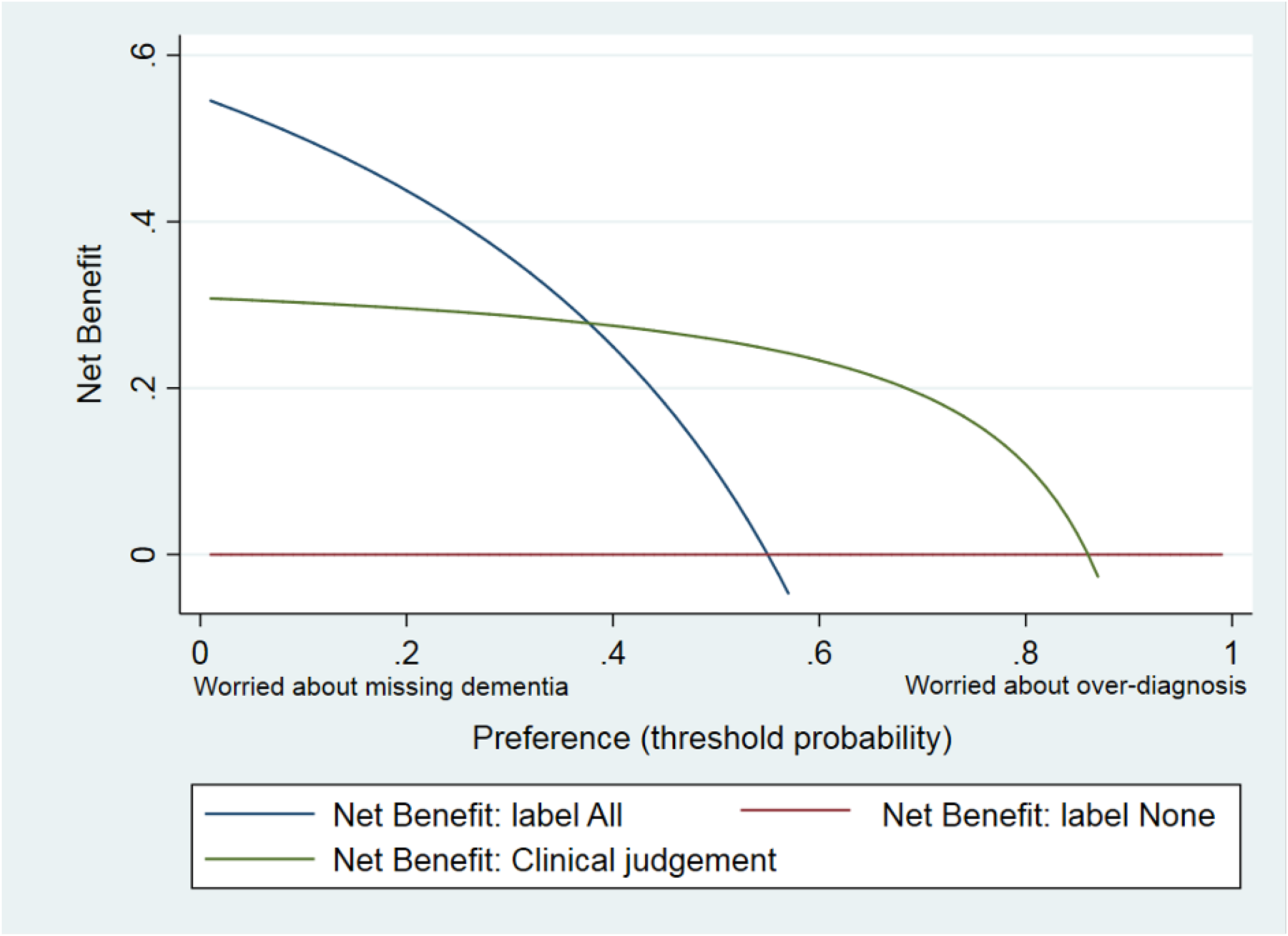
Decision curve showing net benefit of clinical judgement for diagnosis of dementia at a range of threshold probabilities

## Discussion

### Summary

From 21 participating GP surgeries, 456 people were referred and 240 were evaluated. Of these, 132 (55%; 95% CI 48% to 61%) had dementia. Clinical judgement as a single test had a LRP of 5 (95% CI 3 to 9) and a LRN of 0.5 (95% CI 0.4 to 0.6) for the target condition dementia. People that GPs judged as having dementia had a total ACE-III score IQR of 60 to 74, and those that they judged as having MCI had a total ACE-III IQR 71 to 89. This compares to published ACE-III thresholds of <82 for dementia (35) and <88 for MCI (35) and suggests that in this study GPs are not being overly restrictive in their judgement for dementia, or liberal in their judgement for CIND.

### Strengths and limitations

The patient selection in the current study closely reflects real world clinical practice in the United Kingdom, with efforts to avoid people being excluded based on language, transport, or appointment availability. Participants were included with a range of GP opinions about the presence of cognitive impairment in people who had presented with symptoms, which means that cognitive problems were one of the problems discussed in the initial GP consultation; typically 2.5 problems are discussed per appointment (36). The index test clinical judgement in this study reflects an average measure of diagnostic accuracy for an estimated 142 whole time equivalent GPs working in different settings (37). Responses indicated that clinical judgement was typically informed by “face to face presentation”. GPs were told they need not use any formal test to inform their judgement and based on previous studies this is likely to be based on rules of thumb (12) and not formal tests (13). The interval between clinical judgement and the reference standard was relatively short, and unlikely to be associated with a significant progression in cognitive impairment (11). Clinical judgement was fully verified against the reference standard for all consenting people who were referred and there was no evidence of selective participation by cognitive status. Follow-up data after six months was obtained, and uncertain cases were adjudicated. An important limitation is that despite providing translation services the population were largely white, native English speakers. In addition, the confidence intervals for our sub-groups are still wide. We excluded people with advanced cognitive impairment that could not consent, so our findings cannot be generalised to that group; though it is likely that GPs would be more sensitive in identifying cognitive impairment at a more advanced stage.

### Comparison with existing literature

Table 3 summarises the features of this study compared to the existing literature (38, 39). A major strength of this study for applicability to practice is that it is one of only two studies to evaluate symptomatic people. Our study has the smallest number undergoing the index test, but is one of only two studies with complete verification by the reference standard. This study has lower sensitivity and higher specificity than the French study (16), but this could be because the other study verified only 26% of people who underwent the index test, or because other studies did not require participants to be symptomatic and consequently had a lower prevalence of dementia (ranging 2% to 29%).

**Table 3.** Summary of seven studies investigating GP judgement for the diagnosis of dementia

|  | Mannheim<br>(50) | Sydney<br>(51) | Hawaii<br>(52) | Antwerp<br>(15) | AgeCoDe<br>(14) | France<br>(16) | This study |
| --- | --- | --- | --- | --- | --- | --- | --- |
| **Participant selection\*** |  |  |  |  |  |  |  |
| Series | C | C | C | C | R | C | C |
| Symptomatic | No | No** | No | No | No | Yes** | Yes |
| **Characteristics of participants** |  |  |  |  |  |  |  |
| Number (index test) | 3721 | 433 | 303 | 1003 | 3242 | 1453 | 240 |
| Mean age (years) | 76 | 85 | 75 | 75 | 80 | 81 | 80 |
| % Female | 70 | 84 | 63 | 63 | 66 | 71 | 47 |
| % with dementia | 29 | 25 | 9 | 2 | 2 | 50 | 55 |
| **Target condition and verification with reference standard** |  |  |  |  |  |  |  |
| Verified N | 407 | 105 | 303 | 101 | 22 | 385 | 240 |
| Verified % | 11 | 24 | 100 | 1 | 70 | 26 | 100 |
| **GP Judgement (%)** |  |  |  |  |  |  |  |
| Not impaired | 36 | 76 | 33 | - | 94 | 48 | 14 |
| Cognitive impairment | 41 | - | - | - | - | - | 40 |
| Dementia | 23 | 19 | 33 | - | 6 | 26 | 36 |
| Uncertain | - | 5 | 33 | - | - | 22 | - |
| **Diagnostic accuracy of clinical judgement for dementia** |  |  |  |  |  |  |  |
| Sensitivity | 91 | 42 | - | 100 | 51 | 73 | 56 |
| Specificity | 76 | 89 | - | 100 | 96 | 58 | 89 |

C Consecutive R Random Symptomatic: symptoms required for participation
% verified = number underwent reference test / number underwent index test %
with dementia = number with dementia / number verified
\*\* Participants were not presenting with symptoms but GPs were asked to maximise the inclusion of people with suspected dementia – not reported

### Implications for Research and/or practice

Diagnosis can be conceptualised as a pragmatic method of classification that is fit for purpose in the clinical setting (40), and GP judgement may often use heuristics (rules of thumb) and system one (non-analytical (41)) cognitive processes. Clinical judgement may be systematically different to formal definitions, just as different formal definitions (generally formulated for research needs) select different groups of people (42). The GP heuristic of dementia in the older adult may be an individual with forgetfulness who also has sensory impairment, limited mobility, multi-morbidity, and needs additional assistance performing activities of daily life (43). It remains to be seen which definition is most useful in practice. Concerns about resources and lack of specialist expertise for GPs to diagnose and manage dementia well have been reported for many years (44), and GPs have been reported to frame dementia care as a specialist activity (45). However, the priority of patients and their kin who are seeking medical input is to get a prompt diagnosis, in an emotionally safe and personalised way (2). Approaches to diagnosis (3, 6, 46) and follow-up (47) have been reported in primary care but regrettably, a well-designed intervention to improve practice was not effective in improving documentation or increasing case identification (8). In this study clinical judgement was more likely to under than over identify dementia. However, instead of training GPs, patients may benefit more from additional practice-based multi-disciplinary dementia case workers (8) to help refine GP classification, especially where GPs clinical judgement is “not dementia”; in England these roles could be provided through Primary Care Networks (48).

## Data Availability

For access to data please contact the corresponding author

## Additional information

### Ethical approval

The National Research Ethics Service Committee London – Bromley (reference 14/LO/2025) gave a favourable ethical opinion on 25 November 2014. NHS Research and Development approvals were granted by Avon Primary Care Research Collaboration on behalf of Bristol, North Somerset and South Gloucestershire clinical commissioning groups. The University of Bristol acted as Sponsor.

## ACKNOWLEDGEMENTS

The authors thank the participants and the staff at participating practices, without whom this work would not have been possible. The staff at the West of England Clinical Research Network arranged for redaction, collection and transport of medical records from general practices. Written in LATEX using zHenriquesLab-StyleBioRxiv.cls available at https://www.overleaf.com/latex/templates/henriqueslab-biorxiv-template/nyprsybwffws

## Supplementary Note 1: STARD dem checklist

Supplementary Note 1: STARDdem checklist
| Section, topic, and item number | STARD checklist item | Page |
| --- | --- | --- |
| 1 | Identify the article as a study of diagnostic accuracy (recommend MeSH heading “sensitivity and specificity”) | 1 |
| 2 | State the research questions or study aims, such as estimating diagnostic accuracy or comparing accuracy between tests or across participant groups | 1 |
| 3 | The study population: the inclusion and exclusion criteria, setting and locations where data were collected. See also item 4 on recruitment and item 5 on sampling | 1 |
| 4 | Participant recruitment: Was recruitment based on presenting symptoms, results from previous tests, or the fact that the participants had received the index tests or the reference standard? See also item 5 on sampling and item 16 on participant loss at each stage of the study | 2 |
| 5 | Participant sampling: Was the study population a consecutive series of participants defined by the selection criteria in items 3 and 4? If not, specify how participants were further selected. See also item 4 on recruitment and item 16 on participant loss | 2 |
| 6 | Data collection: Was data collection planned before the index test and reference standard were performed (prospective study) or after (retrospective study)? | 2 |
| 7 | The reference standard and its rationale | 2 |
| 8 | Technical specifications of materials and methods involved including how and when measurements were taken, and/or cite references for index tests and reference standard. See also item 10 concerning the person(s) executing the tests | 2 |
| 9 | Definition of and rationale for the units, cutoffs, and/or categories of the results of the index tests and the reference standard | 2 |
| 10 | The number, training, and expertise of the persons executing and reading the index tests and the reference standard. See also item 8 | 2 |
| 11 | Whether or not the readers of the index tests and reference standard were blinded (masked) to the results of the other test and describe any other clinical information available to the readers. See also item 7 | 2 |
| 12 | Methods for calculating or comparing measures of diagnostic accuracy, and the statistical methods used to quantify uncertainty (e.g., 95% confidence intervals) | 2 |
| 13 | Methods for calculating test reproducibility, if done | NA |
| 14 | When study was performed, including beginning and end dates of recruitment | 3 |
| 15 | Clinical and demographic characteristics of the study population (at least information on age, sex, spectrum of presenting symptoms). See also item 18 | 6 |
| 16 | The number of participants satisfying the criteria for inclusion who did or did not undergo the index tests and/or the reference standard; describe why participants failed to undergo either test (a flow diagram is strongly recommended). See also items 3–5 | 7 |
| 17 | Time interval between the index tests and the reference standard, and any treatment administered in between | 3 |
| 18 | Distribution of severity of disease (define criteria) in those with the target condition; other diagnoses in participants without the target condition | 6 |
| 19 | A cross-tabulation of the results of the index tests (including indeterminate and missing results) by the results of the reference standard; for continuous results, the distribution of the test results by the results of the reference standard | 6 |
| 20 | Any adverse events from performing the index tests or the reference standard | 3 |
| 21 | Estimates of diagnostic accuracy and measures of statistical uncertainty (e.g., 95% confidence intervals). See also item 12 | 8 |
| 22 | How indeterminate results, missing data, and outliers of the index tests were handled | 3 |
| 23 | Estimates of variability of diagnostic accuracy between subgroups of participants, readers, or centers, if done | 8 |
| 24 | Estimates of test reproducibility, if done. See also item 13 | NA |
| 25 | Discuss the clinical applicability of the study findings | 3-4 |

